# Caution: The clinical characteristics of COVID-19 patients at admission are changing

**DOI:** 10.1101/2020.03.03.20030833

**Authors:** Zhaowei Chen, Jijia Hu, Zongwei Zhang, Shan Jiang, Tao Wang, Zhengli Shi, Zhan Zhang

**Affiliations:** Department of Nephrology, Renmin Hospital of Wuhan University, Wuhan 430060, China; Department of Dermatology, Renmin Hospital of Wuhan University, Wuhan 430060, China; Department II of Respiratory Disease and Intensive Care, Renmin Hospital of Wuhan University, Wuhan 430060, China; CAS Key Laboratory of Special Pathogens, Wuhan Institute of Virology, Center for Biosafety Mega-Science, Chinese Academy of Sciences, Wuhan 430060, China

**Author notes:** Corresponding author: Zhan Zhang, Department II of Respiratory Disease and Intensive Care, Renmin Hospital of Wuhan University, Wuhan 430060, China. These authors contributed equally to this work (Zhaowei Chen, Jijia Hu, Zongwei Zhang).

**Keywords:** Coronavirus disease-19 (COVID-19), Clinical characteristics

## Abstract

**Background:** With the emergence of 4^rd^ generation transmission, the prevention and treatment of the novel coronavirus disease 2019 (COVID-19) has entered a new period. We aimed to report several changes in the clinical characteristics at admission of patients with COVID-19.

**Methods:** Clinical records and laboratory results of patients suffering from COVID-19 were retrospectively reviewed and matched with the admission dates to analyze the changes in characteristics at the onset of illness.

**Results:** Of the 89 affected patients, 31 [34.8%] patients were admitted from January 16 to 22, and 58 [65.2%] were admitted from January 23 to 29. Patients were admitted with more systemic symptoms, such as fever (21 [67.7%] of 31), fatigue (13 [41.9%] of 31), and myalgia (7 [22.6%] of 31), before January 23. More patients (10 [32.3%] of 31) admitted before January 23 had a small amount of sputum production compared with a smaller proportion (4 [6.9%] of 58) of the patients admitted after January 23. Other symptoms, such as cough, nausea, diarrhea, and chest tightness, were not significantly different between the two groups. In addition, the group admitted before January 23 had a larger proportion of patients with reduced lymphocyte (13 [54.2%] of 24), CD3 (11 [54.4%] of 21), and CD8 (9 [42.9%] of 21) counts and elevated serum amyloid A (SAA, 18 [75%] of 24).

**Conclusions:** The initial symptoms of recently infected patients seem more insidious, indicating that the new coronavirus may gradually evolve into a virus similar to influenza and latent in asymptomatic carriers for a long time.

**Summary:** Compared with the cases admitted earlier, more hidden initial symptoms and improved immune system disorders appeared in COVID-19 patients infected recently.

## Introduction

Since December 2019, there has been an outbreak of large-scale pneumonia cases of unknown cause in Wuhan, China.^1-2^ RT-PCR analyses and electron microscope observations detected a novel coronavirus named SARS-CoV-2 (formerly known as 2019-nCoV).^3^ Coronavirus disease 2019 (COVID-19), which is caused by SARS-CoV-2, has been confirmed to have obvious person-to-person transmission among close contacts.^4-5^ As of February 25, 2020, more than 80,000 confirmed cases and a total of 2700 deaths have been identified globally.^6^ As the epidemic spreads to many countries, COVID-19 poses a severe threat to global health. In the early stage of the epidemic, the most common clinical characteristics of patients with COVID-19 at the onset of illness manifested as fever, cough, and myalgia or fatigue, with or without sputum production and diarrhea.^7^ Therefore, detecting and confirming infected cases in a timely manner and applying scientific treatment as soon as possible is extremely essential to reduce the mortality of COVID-19. Since the emergence of 3^th^ and even 4^rd^-generation transmission^4^ and signs of asymptomatic carrier transmission of COVID-19,^8^ the prevention and control of COVID-19 has entered a new period. Consequently, it is urgent to carefully observe the changes in patients’ initial symptoms. As the designated hospital for patients with severe infection, we have been receiving COVID-19 patients in the newly opened unit since January 16. After the government announced a state of emergency and closed access to Wuhan city on January 23, the number of suspected patients who came to our unit increased rapidly. As of January 29, we received 89 patients confirmed with COVID-19. Surprisingly, during the consultation, we found that the clinical characteristics of patients admitted after January 23 began to differ from those of patients admitted before, which brings new challenges for the diagnosis of suspected patients. To prevent and control the epidemic scientifically, this study aimed to describe the clinical and laboratory characteristics of COVID-19 patients from the perspective of clinical doctors and to compare the initial clinical features between patients infected in different periods.

## Methods

### Study design and patients

The research protocol was reviewed and approved by the Ethics Committee at Renmin Hospital of Wuhan University (Wuhan, China). All research procedures adhered to the tenets of the Declaration of Helsinki. The design of the study was analytical, observational and retrospective. Diagnosis of COVID-19 was based on the New Coronavirus Pneumonia Prevention and Control Program (2nd edition) of China and was indicated by suspected symptoms, chest CT results and SARS-CoV-2 positivity by use of quantitative RT-PCR. We reviewed clinical charts, nursing records, and laboratory findings for 89 patients admitted from January 16, 2020, to January 29, 2020, who had confirmed diagnoses of COVID-19. Patients were assigned into two groups according to their admission date to analyze the differences in patients’ features at admission over time.

### Data collection

The epidemiological data and clinical symptoms of the patients were collected by revising the clinical status (routine blood, CRP, SAA and CD3, CD4, CD8 cells) on the first day of admission from the medical records. Due to incomplete tests for some patients, some types of laboratory results are missing. Three researchers reviewed the data collection forms to double check the data independently.

### Statistical analysis

Categorical variables are described with frequencies and percentages. Quantitative variables are described with the mean (SD) or, if they were not normally distributed, the median. Comparisons of quantitative variables between groups were performed using the independent-sample t test or Wilcoxon rank sum test. Categorical variables were expressed as numbers (%) and compared by χ^2^ test or Fisher’s exact test between the first group and the second group. A two-sided p-value less than 0.05 was considered statistically significant. Statistical analyses were performed using SPSS, version 21.0.

## Results

By January 29, 2020, 89 patients were identified as having COVID-19 and admitted to our hospital. As shown in Table 1, patients were assigned into two groups according to their admission date. Thirty-one [34.8%] of the infected patients were admitted to the hospital from January 16 to 22 (before Jan. 23, first group), and 58 [65.2%] were admitted from January 23 to 29 (after Jan. 23, second group). There was no significant difference in age distribution between the two groups of patients (Table 1). Most of the infected patients were women (45 [77.6%] of 58) in the second group but there were fewer female patients (14 [45.2%] of 31) in the first group (Table 1). The result in the second group is not similar to that from previous studies.^7, 9^ We speculate that this may be related to the small sample size, since there is no evidence that women are more vulnerable to infection as the COVID-19 spreads.

**Table 1:** Baseline characteristics of patients with COVID-19.

| Characteristics | Group 1 | Group 2 |
| --- | --- | --- |
| Age, years, mean (SD) | 33.32 (6.55) <sup>a</sup> | 33.55 (11.73) |
| Sex |  |  |
| Female | 14 (45.2%) | 45 (77.6%) |
| Male | 17 (54.8%) | 13 (22.4%) |
| Admission time | Jan 16 to Jan 22 | Jan 23 to Jan 29 |
| Total patients | 31 | 58 |
| Data are median (SD) or n/N (%), where N is the total number of patients with available data. p values comparing group 1 and group 2 are from independent-sample t test. <sup>a</sup>There is no significant difference between group 1 and group 2. |  |  |

In terms of the initial symptoms, more symptoms appeared in patients from the first group (Table 2), such as fever (21 [67.7%] of 31), fatigue (13 [41.9%] of 31), and myalgia (7 [22.6%] of 31). Several patients (10 [32.3%] of 31) admitted before January 23 had a small amount of sputum production, but only a few people in the second group had this symptom (4 [6.9%] of 58). Other symptoms, such as cough, nausea, diarrhea, and chest tightness, were not significantly different between the two groups. For hematology, the first group had a larger proportion of patients with reduced lymphocyte (13 [54.2%] of 24), CD3 (11 [54.4%] of 21), and CD8 (9 [42.9%] of 21) counts but elevated SAA (18 [75%] of 24) in peripheral blood. Other data, such as CRP, did not show obvious differences between the two groups, suggesting that patients admitted earlier may have had more severe infection and immune disorders (Table 3).

**Table 2:** Comparison of clinical symptoms between two groups.

| Symptom | Group 1 | Group 2 | $\chi^2$ Value | P Value |
| --- | --- | --- | --- | --- |
| Cough | 16 (51.6%) | 24 (41.4%) | 0.491 | 0.483 |
| Sputum | 10 (32.3%)<sup>a</sup> | 4 (6.9%) | 7.983 | 0.005 |
| Fever | 21 (67.7%) | 19 (32.8%) | 9.992 | 0.002 |
| Fatigue | 13 (41.9%) | 8 (13.8%) | 8.875 | 0.003 |
| Myalgia | 7 (22.6%) | 3 (5.2%) | 4.517 | 0.034 |
| Pharyngalgia | 2(6.5%) | 8(13.8%) | 0.480 | 0.489 |
| Nasal symptom | 1 (3.2%) | 0 | 0.000 | 0.348 |
| Diarrhoea | 2(6.5%) | 3 (5.2%) | 0.000 | 1.000 |
| Nausea | 2 (3.2%) | 0 | 0.000 | 0.119 |
| Chest tightness | 5 (16.1%) | 2 (3.4%) | 2.904 | 0.088 |

Data are n/N (%), and N is the total number of patients with available data. p values comparing two groups are from $\chi^2$ test, <sup>a</sup>8 out of 10 patients presented as a small amount of sputum.

**Table 3:** Comparison of blood indicators between two groups.

|  | **All patients**<br>**(N=58)** | **Group 1 (N=24)** | **Group 2**<br>**(N=34)** | **P value** |
| --- | --- | --- | --- | --- |
| WBC, $\times 10^9/L$ | 4.93 (1.52) | 4.39 (1.52) | 5.31 (1.94) | 0.057 |
| <3.5 | 22/58 (37.9%) | 7/24 (29.2%) | 4/34 (11.8%) | 0.161 |
| 3.5-9.5 | 35/58 (60.3%) | 17/24 (70.8%) | 29/34 (85.3%) |  |
| >9.5 | 1/58 (1.7%) | 0 | 1/34 (2.9%) |  |
| LYM, $\times 10^9/L$ | 1.38 (0.43) | 1.19 (0.39) | 1.52 (0.41) | 0.003 |
| <1.1 | 18/58 (31.0%) | 13/24 (54.2%) | 5/34 (14.7%) | 0.001 |
| 1.1-3.2 | 40/58 (69.0%) | 11/24 (45.8%) | 29/34 (85.3%) |  |
| >3.2 | 0 | 0 | 0 |  |
| Mono, $\times 10^9/L$ | 0.49 (0.20) | 0.48 (0.24) | 0.49 (0.17) | 0.859 |
| <0.1 | 0 | 0 | 0 | 0.462 |
| 0.1-0.6 | 43/58 (74.1%) | 19/24 (79.2%) | 24/34 (70.6%) |  |
| >0.6 | 15/58 (25.9%) | 5/24 (20.8%) | 10/34 (29.4%) |  |
| Neu, $\times 10^9/L$ | 2.99 (1.50) | 2.69 (1.31) | 3.19 (1.60) | 0.219 |
| <1.8 | 12/57 (21.1%) | 5/23 (21.7%) | 7/34 (20.6%) | 0.347 |
| 1.8-6.3 | 43/57 (75.4%) | 18/23 (78.3%) | 25/34 (73.5%) |  |
| >6.3 | 2/57 (3.5%) | 0 | 2/34 (5.9%) |  |
| CRP, mg/L |  |  |  |  |
| <10 | 46/58 (79.3%) | 18/24 (75.0%) | 28/34 (82.3%) | 0.725 |
| $\geq 10$ | 12/58 (20.7%) | 6/24 (25.0%) | 6/34 (17.6%) | |
| SAA,mg/L |  |  |  |  |
| <10 | 24/58 (41.4%) | 6/24 (25.0%) | 18/34 (52.9%) | 0.033 |
| $\geq 10$ | 34/58 (58.6%) | 18/24 (75.0%) | 16/34 (47.1%) | |
| CD3, n/$\mu L$ | 864.19 (314.23) | 787.62 (327.45) | 971.40<br>(269.40) | 0.084 |
| <723 | 13/36 (36.1%) | 11/21(52.4%) | 2/15 (13.3%) | 0.016 |
| 723-2737 | 23/36 (63.9%) | 10/21 (47.6%) | 13/15 (86.7%) |  |
| >2737 | 0 | 0 | 0 |  |
| CD4, n/μL | 473.44 (196.80) | 427.86 (183.02) | 537.27<br>(203.71) | 0.101 |
| <404 | 18/36 (50.0%) | 12/21 (57.1%) | 4/15 (26.7%) | 0.070 |
| 404-1612 | 18/36 (50.0%) | 9/21 (42.9%) | 11/15 (73.3%) |  |
| >1612 | 0 | 0 | 0 |  |
| CD8, n/μL | 324.28 (153.84) | 313.14 (172.10) | 339.87<br>(128.10) | 0.614 |
| <220 | 24/36 (66.7%) | 9/21 (42.9%) | 2/15 (13.3%) | 0.008 |
| 220-1129 | 12/36 (33.3%) | 12/21 (57.1%) | 13/15 (86.7%) |  |
| >1129 | 0 | 0 | 0 |  |

Data are median (SD) or n/N (%), where N is the total number of patients with available data. *p* values comparing group 1 and the group 2 are from $\chi^2$ , Fisher's exact test, independent-sample T test or Wilcoxon rank sum test. Abbreviations: WBC, white blood cell; LYM, lymphocyte; Mono, monocyte; Neu, neutrophil; CRP, C-Reactive Protein; SAA, Serum Amyloid A; CD3, CD3 lymphocyte; CD4, CD4 lymphocyte; CD8, CD8 lymphocyte.

## Discussion

We report here the admission characteristics of 89 patients with confirmed COVID-19. All patients were admitted to the designated hospital in Wuhan from January 16 to January 29, 2020. As is known, patients and health-care workers can make a preliminary diagnosis of infectious diseases by accurately grasping the initial symptoms, which is extremely crucial for screening suspicious cases. In the early stage of SARS-CoV-2 transmission, the clinical features of patients with COVID-19 at symptom onset mostly manifest as fever, dry cough, myalgia and fatigue, with or without diarrhea.^7, 9^ However, based on the characteristics of virus mutations and generational transmission, infected patients in different periods may have diverse admission characteristics. Since the 3^rd^ and 4^rd^-generation transmission and even asymptomatic carriers of SARS-CoV-2 have appeared,^4, 8^ it is urgent to distinguish the differences in clinical manifestations of COVID-19 patients at admission during diverse infection periods.

In this study, we observed a small proportion of female patients admitted before January 23, considering that the sample size is not large, it is difficult to account for this phenomenon. However, surprisingly, there were several differences in admission symptoms between the two groups. Some common systemic symptoms of COVID-19, such as fever, fatigue, sputum and myalgia, were more prominent in the patients admitted before January 23, but they were more insidious in patients admitted later. However, there is still no convincing evidence to predict whether the transmission and pathogenicity of SARS-CoV-2 will weaken during transmission. As the “close relatives” of SARS-CoV-2, previous studies have shown that the interpersonal transmission of SARS-CoV and MERS-CoV were constantly changing. Therefore, the clinical manifestations of COVID-19 in different periods should be observed in detail. Research has confirmed that SARS-CoV-2 infects host cells by binding to angiotensin converting enzyme II (ACE2),^10-12^ and ACE2 is highly expressed in nasopharyngeal cells.^13^ More importantly, current research detected a high viral load in the nasopharynx soon after symptom onset.^14^ Theoretically, the nasopharynx is the first virus-infected organ, but through observation, we found that infected individuals rarely show upper respiratory symptoms early in infection, and throat dryness only presented in a few patients. Research and clinical findings seem to suggest that SARS-CoV-2 may be colonized in the nasopharynx but has failed to be recognized in the early stage. As a result, SARS-CoV-2 could hardly be removed by self-cleaning responses such as sneeze and runny and could not be cleared immediately by the local immune system; thus, the initial symptoms of COVID-19 are insidious. Our previous study also indicated that clinical symptoms of COVID-19 deviate from the CT results, and the outcome follows the clinical features.^15^ This further illustrates the critical need to correctly identify the symptoms of COVID-19 at admission. Recently, possible asymptomatic carrier transmission has been reported.^8^ All of these results indicated that the less obvious respiratory symptoms in early infected patients may be due to the existence of pathogenic latency of SARS-CoV-2.

In addition, our study found that a larger proportion of the patients admitted earlier had decreased lymphocyte, CD3 and CD8 T cell counts in the peripheral blood. Reduced CD3 and CD8 T cells counts in the context of excessive activation of these cells have been shown to contribute to the severity and mortality of COVID-19 by causing severe immune damage. ^16^ The contrasting elevated SAA also reflects the infection levels of patients in various periods. Overall, changes in symptoms and laboratory results at admission reflect that SARS-CoV-2 may be undergoing a certain degree of modification, and infection in patients seems to be more insidious. Therefore, we propose that it may be of great significance in guiding the diagnosis and prevention of COVID-19 in the future if respiratory samples, especially lower respiratory samples, could be collected to detect the content of ACE2 and SARS-CoV-2 in patients infected at different periods.

More notably, as of now, 3 unique patients have appeared in our unit. All of them reached the discharge standard (SARS-CoV-2 negativity detected on different days), but a re-examination of their throat swabs showed positive infection days after discharge. One of them was the first confirmed medical staff in our hospital, and this patient has been isolated for more than 20 days after reaching the discharge standard; the remaining 2 patients were isolated 14 days after discharge. These data indicate that the current knowledge of COVID-19 is still insufficient. As the generations of transmissions increase, the clinical manifestations of patients with COVID-19 are changing. The presence of asymptomatic infection also suggests that SARS-CoV-2 may gradually evolve into a virus similar to influenza,^17^ or it may be latent in humans for a long time. However, the limitation of this article is that the number of cases is small so that there may be bias.

## Data Availability

All data referred to in the manuscript are available from the corresponding author by request.

## Author contributions

ZZ designed this study and major in the clinical management of patients, data collection, data analysis, and writing of the first draft. ZC improved the data analysis and the draft. JH and ZZ finished the manuscript and data interpretation. SJ had roles in clinical management of patients. ZS had roles in data analysis and data interpretation, TW helped the data collection. All authors reviewed and approved the final version of the manuscript.

## Acknowledgments

Respectful thanks to Professor Jingping Yuan and her team members Honglin Yan, Dandan Yan for their selfless help. Best wishes to all patients and their families in this study. Mourn all those who died in this disaster.

## Data Availability Statement

All data referred to in the manuscript are available from the corresponding author by request.

